# An intuitive sampling framework for setting-specific decision-making in soil-transmitted helminthiasis control programs

**DOI:** 10.64898/2026.02.11.26346062

**Authors:** Adama Kazienga, Bruno Levecke, Sake J. de Vlas, Luc E. Coffeng

## Abstract

**Background:** We recently developed a general egg count framework to support cost-efficient survey design choices to inform soil-transmitted helminthiasis (STH) control programs. Yet, the interpretation and the application was not always intuitive for program managers.

**Methods:** We first adapted the existing framework to make the interpretation of risks of incorrect decision making more intuitive and to allow for prior information. Then, we assessed the impact of the allowable risk of incorrect decision-making and prior information on the required sample size. Finally, we determined the most cost-efficient survey design to inform the decisions (i) to switch to an event-based deworming program, and (ii) to declare STH eliminated as a public health problem (EPHP).

**Principal findings:** The required sample sizes increased when the allowable risk of incorrect decision reduced and when the mean prior approached the program prevalence threshold. For the decisions to switch to event-based deworming and to declare EPHP, we found that duplicate Kato-Katz thick smears on a single stool sample was the most cost-efficient survey design, particularly when particularly when accounting for the added benefits of the free internal quality control. The required sample size for these survey designs varied between program targets and STH species. When aiming to have one sample size that fits all STHs, we recommend sampling 6 schools and 56 children per school for decisions on switching to event-based control programs and 11 schools (74 children per school) for the decision to declare EPHP.

**Conclusions/significance:** We developed an intuitive sampling framework for setting-specific decision-making in STH control programs. We identified the most cost-efficient survey designs for critical program decisions, but these are based on subjective but reasonable choices regarding the risk of incorrect decision making. Reaching consensus within the STH community on acceptable levels of risk is crucial to further support evidence-based decision-making.

**Author summary:** We recently developed a general computer simulation framework to support cost-efficient survey design choices for the control of intestinal worms. However, its interpretation was not always intuitive and it did not allow incorporation of prior knowledge on the prevalence of infections that programs might have. In this study, we adapted our framework to make the risks of incorrect decision-making more intuitive to interpret and to incorporate prior information on worm prevalence. We then quantified how different risk tolerances and prior prevalence assumptions affected required survey designs. Using this framework, we then identified the most cost-efficient survey designs for two key program decisions: switching to event-based deworming and declaring elimination of intestinal worms as a public health problem. We found that lower tolerance for incorrect decisions and greater uncertainty around prior prevalence substantially increase required sample sizes. Across the different program decisions and worm species, examining duplicate Kato–Katz thick smears from a single stool sample was consistently the most cost-efficient design, with the added benefit of internal quality control. Our results provide practical guidance for designing surveys tailored to local settings and highlight the importance of reaching consensus on acceptable levels of decision-making risk to support evidence-based STH control.

## Introduction

Soil-transmitted helminths (STHs; *Ascaris lumbricoides, Trichuris trichiura*, and two hookworm species *Necator americanus* and *Ancylostoma duodenale*) are intestinal worms that are transmitted through the uptake of infectious life stages from the environment (often soil). In 2021, an estimated 1.5 billion people were infected worldwide [1,2], the vast majority living in resource-constrained countries, where lack of sanitation and limited access to clean water maintain transmission. To reduce STH-attributable morbidity (1.38 million disability life years lost in 2021 [3]), the World Health Organization (WHO) recommends large-scale deworming programs during which entire at-risk groups (school-age (SAC) and pre-school-age children and women of reproductive age) are periodically dewormed – the so-called preventive chemotherapy (PC) programs. In 2021 alone, more than 500 million children and 99 million women of reproductive age in STH-endemic areas received a treatment, resulting in a respective program coverage of 55% and 40% of the target populations at risk [4]. Encouraged by the reported reduction in STH global burden of 38% from 2012 to 2021 [3], WHO recently released its new roadmap for the period 2021-2030. While the targets in the previous road map (2012-2020) focused on increasing the program coverage only [5], the two most important targets are now the reduction in the tablets needed in STH deworming programs and the elimination of STHs as a public health problem (EPHP), which is defined as a prevalence of moderate-to-heavy intensity (MHI) infections of less than 2% [2].

To assess the impact of control strategies and to ensure progress towards the WHO 2030 targets for STHs, periodic follow-up surveys are crucial. These surveys, referred to as monitoring and evaluation (M&E), serve to inform decisions on whether continuing or scaling down PC is justified. To support this decision-making, the WHO released guidance based on the prevalence thresholds of any STH infections (i.e., 2%, 10%, 20%, and 50%). According to this guidance, the frequency of deworming should decrease with decreasing STH prevalence, with any STH prevalence below 2% triggering so-called event-based PC. This event-based strategy refers to the administration of deworming medication during specific events such as immunization visits for preschool-age children, school enrolment or graduation for SAC, and antenatal care visits for women of reproductive age [6]. To determine the prevalence of STH infections during the M&E, the WHO recommends screening stool samples from 250 SAC across 5 schools (50 children per school), collecting one stool sample per child, and examining one Kato-Katz (KK) thick smear slide per sample [7–10].

Importantly, the process of decision-making during M&E always carries the risk of making a wrong decision: switching to event-based PC or declaring EPHP too early (referred to as “undertreatment”) or too late (“overtreatment”). The magnitude of these risks is determined by the accuracy of the survey strategy, which depends on sample size, specifically the number of schools surveyed, and the number of children sampled per school. Note that accuracy also depends on the diagnostic approach, including the number of stool samples and the number of KK thick smears per sample. However, improving survey accuracy through larger sample sizes and more intensive diagnostic efforts entails significant financial and logistical investments. As such, there is a need to balance operational costs and feasibility of surveys with the maximum allowable risks of under- and overtreatment. In our efforts to support cost-efficient survey design choices, we previously developed a simulation framework around a lot quality assurance sampling (LQAS) strategy that explicitly captures these risks of under-and overtreatment [11]. An important limitation of this framework is that the interpretation of the risks of incorrect decision-making is not always intuitive for program managers. Moreover, the existing framework does not incorporate prior knowledge regarding the true prevalence, limiting the tailoring of survey design to local epidemiological contexts.

In this study, we adapted the existing LQAS framework to make the interpretation of risks of incorrect decision-making more intuitive for program managers. Next, we explored the impact of acceptable levels of risk of over- and undertreatment and prior knowledge around true prevalence on survey design choices. Finally, we identified the most cost-efficient survey design for STH control programs.

## Methods

### Ethics statement

We used a dataset from a study in Ethiopia, which has already been published elsewhere[12], to quantify sources of variation in egg count. This study was designed for the national mapping of soil-transmitted helminth and schistosome infections in Ethiopia. The study protocol was reviewed and approved by the Ethiopian Public Health Institute Scientific and Ethical Review Office (reference number SERO-128-4-2005) and the Institutional Review Board of Imperial College London (reference number ICREC_8_2_2). Verbal consent was obtained from the parents or guardian, and the school head provided written consent on behalf them. In addition, students provided verbal consent to be included in the survey.

### Overview

We revised the existing LQAS framework [11] in two steps. In the first step, we expanded the framework to make the interpretation of the risk of incorrect decision-making more intuitive for program managers. As this expansion required prior information on the expected true prevalence, we initially assumed a uniform prior knowledge and compared this revised approach conceptually to the existing framework. In the second step, we further extended the framework to allow for more realistic levels of prior information regarding the expected true prevalence based on the mean prior and the associated uncertainty. Using this revised framework, we built a database of simulations representing different scenarios of survey design (number of schools, children per school, stool samples per child, and KK thick smears per stool sample). Finally, we analysed the database (i) to assess the impact of prior knowledge and the risk of incorrect decisions on both the required sample size and total survey costs, and (ii) to identify the most cost-efficient survey design for STH control programs.

### Expansion of the existing LQAS framework

We expanded our existing framework to make the interpretation of the risk of incorrect decision-making more intuitive for program managers. Specifically, we defined the risk of overtreatment (= *O*) when the true underlying prevalence is below the program prevalence threshold and the risk of undertreatment (= *U*) when the true underlying prevalence is at or above this threshold. Their formulation is as follows:

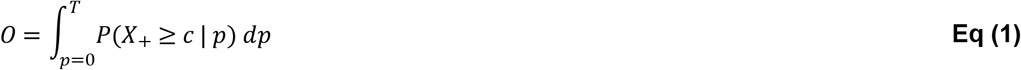

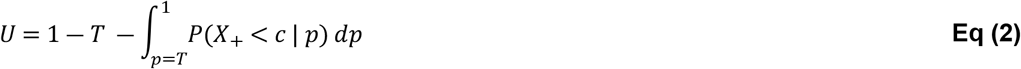

Here, *p* is the true but unknown prevalence of infection, *T* represents the program prevalence threshold (e.g., 2%), *X*_+_ is the total number of positive test results (both true and false positive test results), and *c* represents the decision cut-off. Note that these equations are conditioned on the number of schools *n*_*schools*_, and the number of children *n*_*children*_ sampled per school, the number of stool samples per child, and the number of repeated KK thick smears per stool sample.

The conceptual difference between the existing and expanded frameworks is illustrated in **Fig 1**, with **Fig 1A** representing the former and **Fig 1B** the latter. Both panels show the probability of continuing PC as a function of the true underlying prevalence for a particular survey design, assuming a program prevalence decision threshold of 2%. In the previous framework, the concept was to ensure that the risk of making an incorrect program decision at two predefined values of true underlying prevalence does not exceed a predefined risk. These values correspond to the highest possible true prevalence below (*LL*: lower limit) and the lowest possible true prevalence above the program prevalence threshold (*UL*: upper limit) that should trigger reliable decision-making [11]. In contrast, the revised framework extends this concept by determining the risk of incorrect decision-making across all possible values of true underlying prevalence. The risk of overtreatment (= *O*) is represented by the area under the curve (in yellow) when the true underlying prevalence is below the program prevalence threshold, and the risk of undertreatment (= *U*) is represented by the area above the curve (in red) when the true underlying prevalence is at or above this threshold.

**Fig 1:**
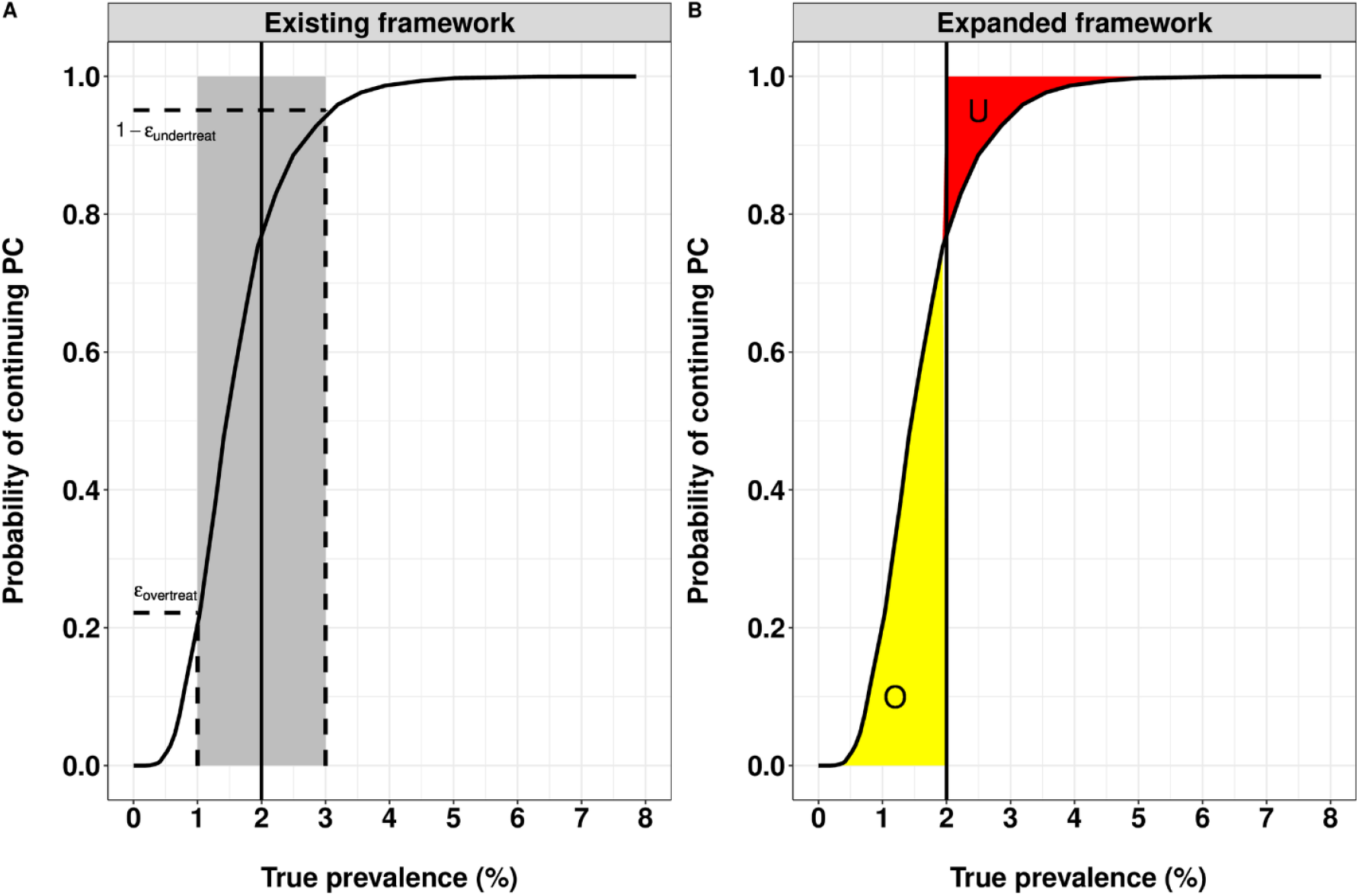
A conceptual comparison of the existing and the expanded framework. This graph illustrates the conceptual difference between the existing framework (**Panel A**) and the expanded framework (**Panel B**) regarding the estimation of the risk of incorrect decision-making. In the existing framework, the risks of making an incorrect program decision (the horizontal dotted lines in **Panel A** represent the achieved risks of overtreatment and the risk of undertreatment) are conditioned on two chosen values of true underlying prevalence (vertical dotted lines in **Panel A**). In contrast, the new framework (**Panel B**) estimates these risks across all range of true underlying prevalence values (the surfaces *O* and *U* represent the risk of over- and undertreatment, respectively), providing more intuitive interpretation. This graph shows the probability of continuing preventive chemotherapy when 396 children across 6 schools were screened for hookworm infections with duplicate Kato-Katz thick smears based on a single stool sample, using a decision cut-off of *c* equal to 9 positive test results. The vertical straight line in both panels indicates the program prevalence threshold *T* = 2%. Note that for Panel B, we assumed no prior knowledge about the true underlying prevalence (i.e., all values between 0% and 100% are weighted equally).

Note that **Eqs (1)** and **(2)** assume that all values of the true underlying prevalence *p* are equally likely before having observed the data (i.e., a uniform prior for *p*). However, this is not reasonable because we are considering to check whether the prevalence is above or under threshold *T*, implying that we believe it to be close to or less than *T*. A program manager might base such a prior expectation of the true prevalence on what they know about the prevalence of infection before the start of PC and the duration and therapeutic coverage of PC, or on recent survey that took place in the same area. Therefore, we further expanded the framework to relax the assumption of a uniform prior for the true prevalence *p*. We describe this prior knowledge with the beta distribution, which is defined in terms of parameters *α* and 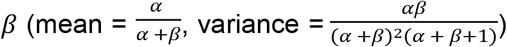 . For this study, we parametrized the beta distribution in terms of the mean *µ* and the degree of certainty *κ* (where *κ* = *α* + *β*). The corresponding shape parameters can be derived as a function of mean *µ* and degree of certainty *κ*, defined as *α* = □*μ* and *β* = (1 ― *μ*) □.

We further assessed the impact of the aforementioned critical parameters on the shape of the beta distribution (**Fig S1**). First, we explored the impact of the degree of certainty κ on the shape of the beta distribution while fixing the mean *μ*. To this end, we first set the mean *µ* to 1% (i.e., half the decision threshold of 2%) while considering degree of certainty *κ* values of 50, 200, and 800 (**Fig S1A**). We noted that when □ is relatively low (e.g., □ = 50), the beta distribution becomes J-shaped, with a high density near zero, reflecting a strong belief that prevalence is low. Note that as □ further decreases, the beta distribution becomes a U-shaped distribution, with density concentrated near 0% and 100%. At moderate □ values (□ = 200), the beta distribution becomes bell-shaped with probability mass below and above the mean *μ*. In contrast, when □ is high (*□* = 800), the density becomes sharply peaked around the mean, approximating a narrow bell curve compared to moderate □ beta distribution shape. Second, we assessed the impact of mean *μ* on the shape of the beta distribution when considering a fixed degree of certainty □ (**Fig S1B**). When fixing □ at 200, the shape of the beta distribution varies with changes in the mean *µ*. At a mean *µ* of 0.5%, the distribution is J-shaped, with high density near zero and a long right tail, reflecting a strong belief that prevalence is very low and under the decision threshold. As the mean *µ* increases from 0.5% to 1%, the distribution becomes bell-shaped, indicating greater concentration around the mean. When the mean *µ* further increases from 1% to 1.5%, the bell-shaped curve becomes more symmetric around the mean. These differences highlight how, even with the same degree of certainty □, the location of the mean *µ* affects the prior belief regarding where the true prevalence lies.

As including prior knowledge impacts the estimation of *O* and *U*, we need to rewrite **Eqs (1)** and **(2)** into:

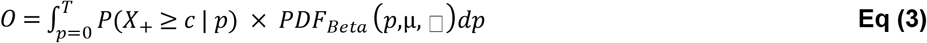

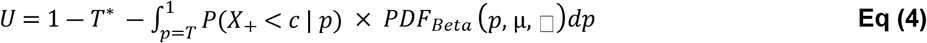

where

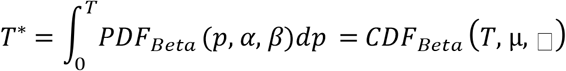

Here, *PDF*_*Beta*_ is the probability distribution function of the beta distribution, and *CDF*_*Beta*_ is the associated cumulative distribution function.

### Build-up of the database of simulations

The database of simulations was generated by following a similar procedure as extensively described in our previously published paper [11] through a structured multi-step process. In **the first step**, we determined the mean number of eggs per gram stool (EPG) at the implementation unit level that corresponds to the true prevalence (*p*) of infection (any intensity and MHI intensity) when deploying single KK thick smear on a single stool sample, accounting for the different sources of variability in egg counts as estimated in previous studies (**Table S1**). For this, we used 150 values of mean EPG as input on a logarithmic grid, resulting in 150 point estimates of true prevalence, varying from 0 to 100% when hypothetically testing 3,000 schools and recruiting 6,000 children per school at the implementation unit-level. **Fig S2** and **S3** illustrate the resulting association between mean EPG and true prevalence of infection (any intensity and MHI) at the implementation unit level, respectively. In **the second step**, we simulated KK thick smear egg counts for stool samples collected from all sampled children in the survey, and we determined the children who tested positive. For this, we let the number of schools (*n*_*schools*_) varies from 3 to 10 with increments of 1, and allowed the number of children per school (*n*_*children*_) to vary from 10 to 100 (a reasonable maximum size of individuals in a school) with increments of 2. As previously, we defined the stool collection and examination procedure as *KK*_*a*×*b*_, where *a* is the number of stool samples per child, and *b* is the number of repeated KK smears per stool sample. Note that both *a* and *b* were allowed to take values 1 or 2 in four distinct stool examination scenarios (*KK*_1×1_, *KK*_1×2_, *KK*_2×1_, and *KK*_2×2_). Among these four scenarios, only three (*KK*_1×2_, *KK*_2×1_, and *KK*_2×2_) allow for internal quality control of the egg counting process, enabling independent verification of results and enhancing diagnostic reliability. This is in contrast to *K*_1×1_, which does not include this component. Using these numbers resulted in 220,800 combinations (4 scenarios of stools examination × 8 values of *n*_*schools*_ × 46 values of *n*_*children*_ × 150 grid points of true prevalence). For each combination, 10,000 Monte Carlo iterations were simulated. In **the third and last step**, we calculated the number of working days required to screen all recruited children for each survey design (*n*_*schools*_, *n*_*children*_, number of stool samples per child, and number of KK thick smears per stool sample) as previously described [11].

### Determining the most cost-efficient survey design

We then evaluated the simulated dataset to identify the most cost-efficient survey for reliably determining whether infection prevalence is below or above the decision threshold. Using the database of simulations, we estimated the total survey costs and the risks of incorrect decision-making for all combinations of survey designs and potential decision cut-offs (*c*). For the total survey costs, we followed a similar procedure that is extensively described in our previously published paper [11]. This cost is based on the number of working days that a field team would need to screen all recruited children and includes the consumables cost (cost to collect and process samples), the personnel cost (per diem for laboratory technicians and nurses), and the travel cost (car rental, driver wage, and gasoline) in an Ethiopian setting [11–13] (**Table S2**). The risk of over- and undertreatment was calculated using **Eqs (3)** and **(4)**. To explore the impact of the prior knowledge, we considered 40 values of *κ* (degree of prior certainty; ranging from 100 to 1,000, equally spaced on the logarithmic scale) and six values for the prior mean *μ* (0.5%, 1%, 1.1%, 1.2%, 1.3%, 1.4%, and 1.5%). The values for the prior mean *μ* were chosen to reflect settings nearing the 2% prevalence threshold, a critical benchmark for switching to event-based PC and for declaring EPHP. Note that for each mean *μ*, we identified the most conservative value of *κ*, defined as the level of prior certainty that yields the largest sample size to ensure that the resulting survey design remains sufficiently powered for reliable decision-making. To also identify cost-efficient survey design choices around the other prevalence thresholds (10%, 20% and 50%) to scale down PC, we considered threshold-specific prior means of 5% for the prevalence threshold of 10%, 15% for the prevalence threshold 20%, and 40% for the prevalence threshold 50%, combined with the most conservative *κ* (i.e, the level prior uncertainty that yields the largest sample size) for each value of the prior mean. Because we aim to provide recommendations that can cover a wide range of scenarios of STH endemicity (single *vs*. multiple co-endemic infections), we also explored whether we could recommend one sample size for all three STHs, with species-specific decision cut-offs to ensure reliable decision-making for each of the three STH.

Finally, we identified the survey designs that allowed for reliable program decision-making (not exceeding the maximum acceptable risk of over- and undertreatment) at the lowest operational cost across different maxima for under- (0.5%, 1%, and 2%) and overtreatment (15%, 20%, and 25%).

## Results

### Illustration of the expanded framework

We first illustrate our expanded framework to determine the most cost-efficient survey design when focusing on event-based PC (prevalence threshold of 2%) in an area where only hookworm infections occur. This illustration highlights four important aspects (**Fig 2)**. First, as expected, the required number of children per school decreases as a function of increasing number of sampled schools (**Fig 2A**). Second, collecting more stool samples per child and preparing more smears per stool sample (*KK*_1×2_, *KK*_2×1_ and *KK*_2×2_ *vs. K K*_1×1_) allows for fewer children to be sampled per school, as expected from increased sampling effort. For instance, when considering a program decision for hookworm and sampling six schools (**Fig 2A**), 68 children per school are required for a *KK*_1×1_ survey design, while this number drops 18% to 29% as more samples and slides per sample are screened per child (*KK*_1×2_: 56, *KK*_2×1_: 58, and *KK*_2×2_: 48). Third, sampling a second stool sample (*KK*_2×1_ and *KK*_2×1_) results in higher total survey costs, even though there is a reduction in the required number of children per school (**Fig 2B**). For example, when sampling six schools, the total survey cost for a *KK*_1×1_ survey design equals 2,951 US$ (*n*_*children*_ = 68), while the survey cost increased to 4,590 US$ (*n*_*children*_ = 58) when deploying a *KK*_2×1_. Fourth, the total survey costs are lower when fewer schools are sampled, as it takes fewer days to complete the survey (**Fig 2B**). However, when the number of sampled schools becomes too low, more children need to be sampled per school, and hence, an extra day is sometimes needed per school, resulting in an increase in total survey cost (e.g., 5 *vs*. 6 schools when deploying *KK*_1×2_; green line in **Fig 2B**). When considering all aspects illustrated by **Fig 2**, *KK*_1×1_ was identified as the most cost-efficient survey design for switching to an event-based PC when focusing on hookworm infections only, as indicated by the black bullet point in **Fig 2B**. However, when considering internal quality control of the egg counting process, *KK*_1×2_ emerged as the most cost-efficient option, as denoted by the open circle in the same figure. These patterns were very similar for the other two STH species as shown in **Fig S4**.

**Fig 2:**
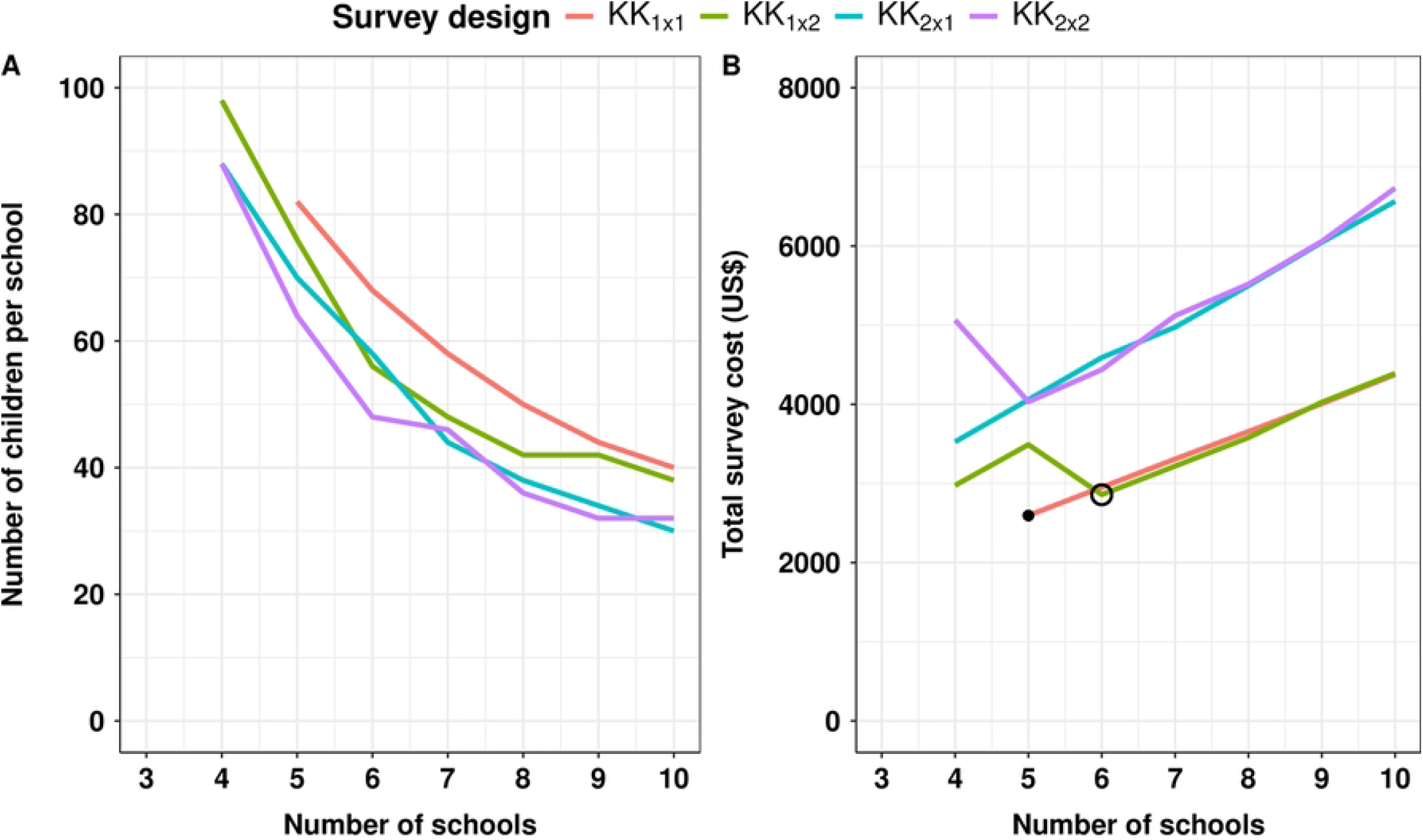
Identification of the most cost-efficient survey design to reliably switch to event-based PC. This figure illustrates the required number of children per school (*n*_*children*_; **Panel A**) and the corresponding total survey cost (*C*_*tot*_; **Panel B**) as a function of the number of sampled schools (*n*_*schools*_) for different survey designs in an area where hookworm is the only or main prevalent STH species. The survey designs (*KK*_*a* × *b*_) varied in the number of stool samples per child (*a*) and the number of Kato-Katz thick smears per sample (*b*). The black bullet point in Panel B indicates the most cost-efficient survey design when preparing a single slide per sample. The open circle indicates the most cost-efficient survey design when preparing two slides per sample, which automatically adds free quality control to the survey design. Note that when a line does not extend further to the left in **Panels A** and **B**, it indicates that the corresponding survey design failed to meet the maximum acceptable level of risk of making an incorrect decision at those lower numbers of sampled schools (e.g., 3 schools).

### Impact of prior knowledge and maximum acceptable risk of incorrect decisions on survey design

Next, we explored the impact of prior knowledge (mean *μ* and degree of certainty *κ*) on the required sample size and total survey cost to make decisions regarding switching to event-based PC (**Fig 3**). We observed two important aspects. First, the relationship between □ and the required total survey cost is concave. The required total survey cost increases as a function of increased prior certainty □, reaches a peak (turning point), and then gradually drops. Accordingly, for each value of mean *μ*, we identified the most conservative □, defined as the value yielding the highest total survey cost (**Table S3)**. The second important observation is that the total survey cost increases as the values of the prior mean approaches the decision threshold when using the same degree of certainty □. For example, when considering the most conservative □ of 200 (**Fig S5**), the corresponding survey cost was 1,142 US$ for 0.5% prior mean, while this was 2,858 US$ for 1% and 7,136 for 1.5% prior mean (**Fig 3B**). This pattern illustrates the rationale behind identifying the most conservative □, defined as the value that yields the maximum required survey cost for a given prior mean, thereby ensuring that the survey design is sufficiently powered.

**Fig 3.**
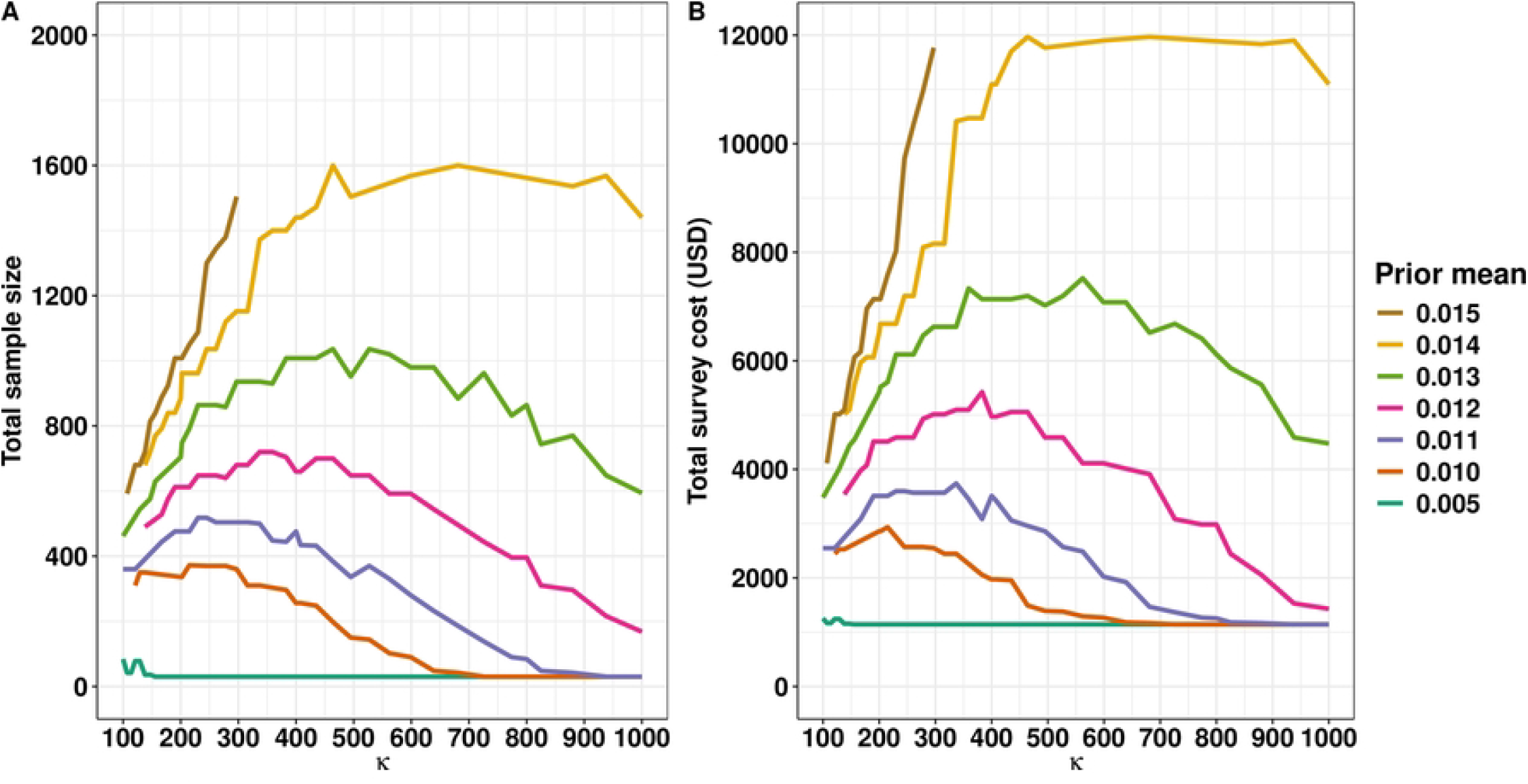
Impact of prior knowledge on the sample size and total survey cost to switch to an event-based PC. This figure represents how prior mean and the degree of certainty □ Influence the sample size (**Panel A**) and the total survey cost (**Panel B**). Here, we set the maximum acceptable risk of undertreating *U* to 1% and the risk of overtreatment (*O*) to 20% and assumed a survey design based on a duplicate Kato-Katz thick smear using a single stool sample (*KK*_1×2_).

We further assessed the impact of the maximum allowable risk of incorrect decision-making on the total survey cost (**Fig 4**). To this end, we first fixed the maximum acceptable risk of overtreatment at 20% (**Fig 4A**) while varying the maximum acceptable risk of undertreatment (0.5%, 1%, and 2%). Next, we fixed the maximum acceptable risk of undertreatment at 1% (**Fig 4B**) and then varied the maximum acceptable risk of overtreatment (15%, 20% and 25%). A key observation in these panels is that minimizing the risk of incorrect decision-making resulted in substantially higher total survey costs.

**Fig 4:**
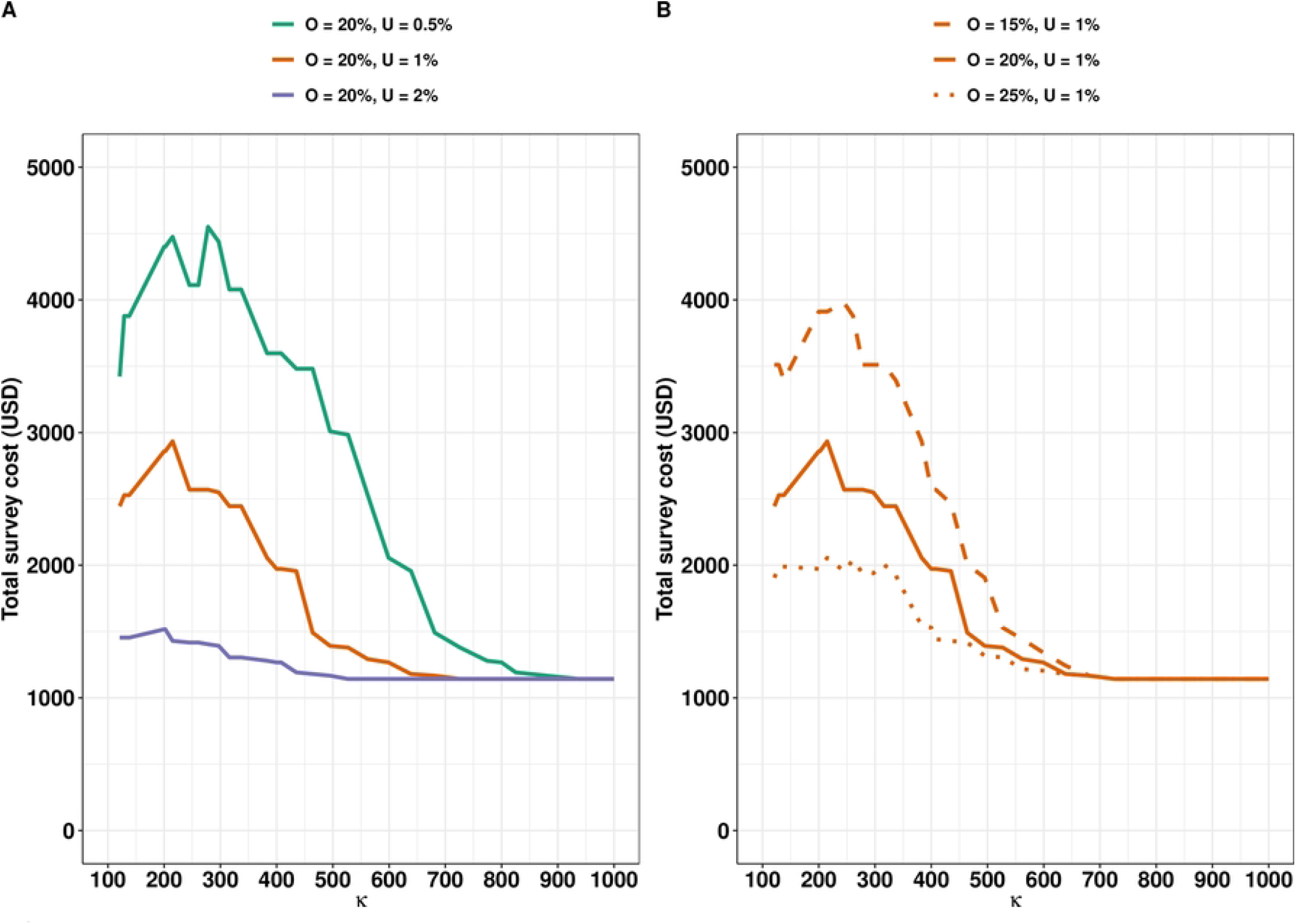
Impact of the maximum acceptable risks of under- and overtreatment on the total survey cost to event-based PC. This figure represents the required total survey cost for different prior certainty levels □ and levels of risk of making an incorrect decision. **Panel A** shows the impact of the risk of undertreatment, and **Panel B** presents the impact of the risk of overtreatment, assuming a prior mean of 1% and a survey design based on a duplicate Kato-Katz thick smear on a single stool sample (*KK*_1×2_). The solid brown line in **Panel A** represents the main analysis, serving as a reference scenario in **Panel B**.

For instance, a reduction in risk of undertreatment from 2% to 1% increased the total survey costs by 89% (1,516 US$ *vs*. 2,858 US$, given *κ* = 200 and a maximum risk of overtreatment of 20%) (**Fig 4A**). When we further reduced the risk of undertreating to 0.5%, the total survey cost further increased by 54% (2,858 US$ *vs*. 4,400 US$). Similar patterns were observed when exploring the impact of the risk overtreatment on the total survey costs (**Fig 4B**).

### Most cost-efficient survey design for STH control programs

#### Switching to event-based PC

To identify the most cost-efficient survey design to make decisions to switch to event-based PC, we fixed the mean *μ* at 1% while the degree of certainty *κ* was set to 200, a combination that we considered a reasonable choice (**Fig S5**). Also, we chose both single and duplicate KK scenarios on a single stool sample, as they were found to be equally cost-efficient for all STHs, as illustrated in **Fig S5. Table 1** subsequently presents the most cost-efficient survey design to switch to event-based PC across the three STH species when deploying single and duplicate KK thick smears on a single stool sample. Notable differences in required sample size (*n*_*schools*_ × *n*_*children*_) across the three STH species were observed, regardless of whether single or duplicate KK was used. For example, when considering a *KK*_1×1_ survey design, the most cost-efficient sample size was 4 schools and 80 children per school (4 x 80) for *Ascaris* infection, 5 x 82 for hookworm infections, and 4 x 100 for *Trichuris* (**Table 1**). When using *KK*_1×2_, the most cost-efficient survey design was 4 x 70 for *Ascaris*, 6 x 56 for hookworm, and 5 x 68 for *Trichuris* (**Table 1**). While *KK*_1×1_ and *KK*_1×2_ offers cost-efficient survey design across the three STH species, *KK*_1×2_ was associated with slightly higher total survey cost. For instance, the total survey cost increased by 10% (2,595 US$ *vs*. 2,858 US$) for hookworm and 21% (2,216 US$ *vs*. 2,507 US$) for *Trichuris*. Despite this modest increase in total survey cost, *KK*_1×2_ allows for the internal quality control of the egg counting, which may improve diagnostic reliability and offset potential misclassification costs.

**Table 1.**
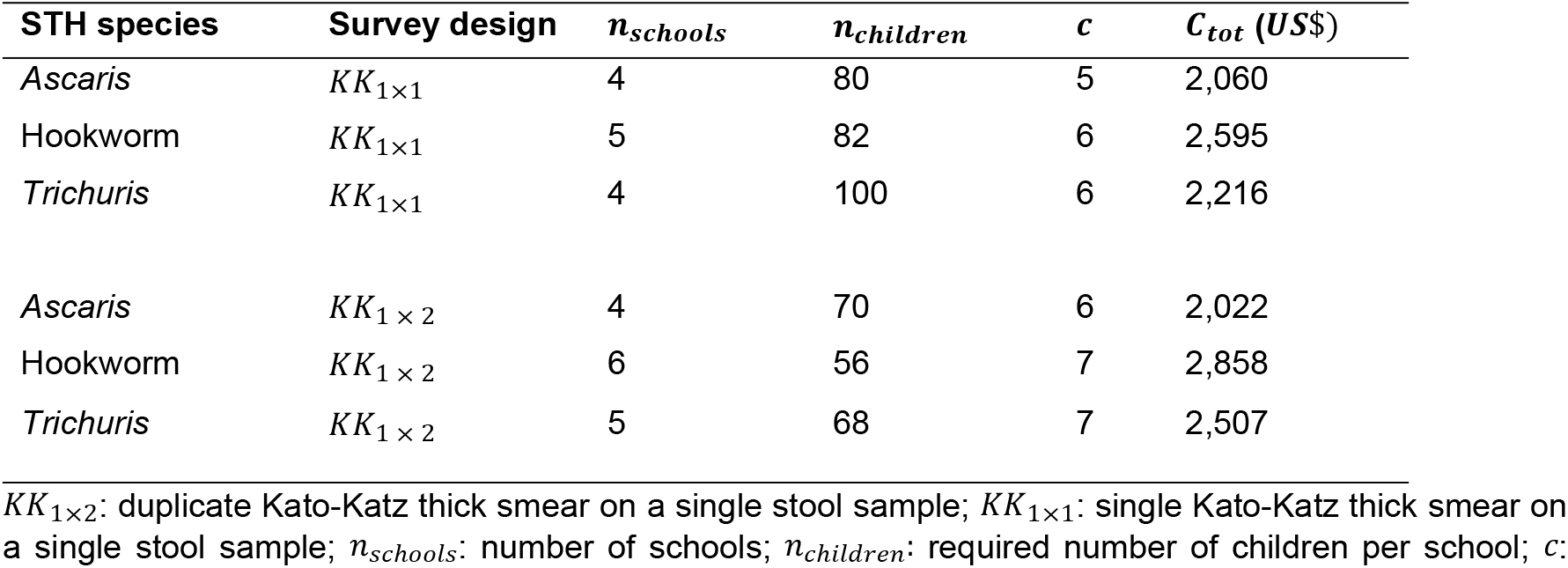
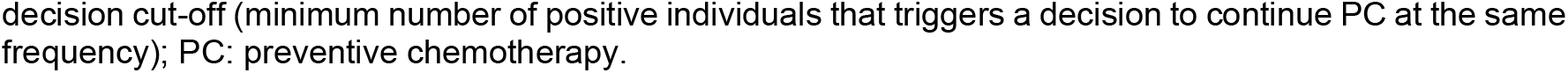
The most cost-efficient survey design to switch to an event-based PC. This table represents the required sample size (*n*_*schools*_ × *n*_*children*_), the decision cut-off *c* and the total survey cost (*C*_*tot*_) for surveys based on screening one stool sample with a single (*KK*_1×1_) and duplicate Kato-Katz thick smear (*K K*_1×2_). We further set the maximum acceptable risk of undertreatment at 1% and the maximum acceptable risk of overtreatment at 20%, a certainty level □ of 200, and a mean *μ* of 1%.

| STH species | Survey design | $n_{schools}$ | $n_{children}$ | $c$ | $C_{tot}$ (US\$) |
| --- | --- | --- | --- | --- | --- |
| *Ascaris* | $KK_{1 \times 1}$ | 4 | 80 | 5 | 2,060 |
| Hookworm | $KK_{1 \times 1}$ | 5 | 82 | 6 | 2,595 |
| *Trichuris* | $KK_{1 \times 1}$ | 4 | 100 | 6 | 2,216 |
| *Ascaris* | $KK_{1 \times 2}$ | 4 | 70 | 6 | 2,022 |
| Hookworm | $KK_{1 \times 2}$ | 6 | 56 | 7 | 2,858 |
| *Trichuris* | $KK_{1 \times 2}$ | 5 | 68 | 7 | 2,507 |

$KK_{1 \times 2}$ : duplicate Kato-Katz thick smear on a single stool sample; $KK_{1 \times 1}$ : single Kato-Katz thick smear on a single stool sample; $n_{schools}$ : number of schools; $n_{children}$ : required number of children per school; $c$ :
decision cut-off (minimum number of positive individuals that triggers a decision to continue PC at the same frequency); PC: preventive chemotherapy.

We further determined the recommended survey design for STH control programs to support switching to event-based PC for areas where the prevalence of mixed STH infections are high. To this end, we considered the largest sample size across all three STH species when deploying *KK*_1×1_ or *KK*_1×2_ (**Tables 1 and S4**) but determined STH-specific corresponding decision cut-offs that ensure reliable decision-making across all STHs. This corresponds to sampling 5 schools (82 children per school) for *KK*_1×1_ and 6 schools (56 children per school) for *KK*_1×2_ for switching to event-based PC or scaling PC. Although cut-offs were determined separately for each STH species, for switching to event-based PC, they converged to 6 for *KK*_1×1_ and 7 for *KK*_1×2_.

#### Declare EPHP

Using the same approach to identify the most cost-efficient survey design to switch to an event-based PC, we determined the most cost-efficient survey designs to declare EPHP for all three STH species when deploying single and duplicate KK thick smear on a single stool sample (**Table S4**). Compared to decisions regarding event-based PC (**Table 1**), we note that the sample size required for declaring EPHP is substantially larger. To recommend a single survey design to declare EPHP for all species, we again adopted the most conservative survey design across the STH species (i.e., the largest sample size) and determined the STH species-specific decision cut-off. As for a switch to event-based PC, this sample size was again dictated by hookworm. As such, to declare EPHP, the required survey size was 11 schools (74 children per school), and decision cut-off was 11 for hookworm and 10 for *Ascaris* and *Trichuris* (**Table S4**).

#### Recommended survey design for other STH control program decisions

Furthermore, we determined the most cost-efficient design to scale down the frequency of PC (e.g., 10%, 20%, and 50% prevalence threshold). Given that the required sample size to make decision around the 2% prevalence threshold remains the largest compared to other thresholds (i.e., 10%, 20%, and 50%) [14], we continued to use 5 x 82 for *KK*_1×1_ and 6 x 56 sample for *KK*_1×2_, and determined the corresponding threshold-specific decision cut-off for each STH separately. The threshold-specific decision cut-offs are summarized in **Table 2** for each of the three STH species. For instance, the decision cut-offs for hookworm were 27 out of 6 x 56 children (*KK*_1×2_.) for the prevalence threshold of 10%, 74 for a 20% prevalence threshold, and 178 for a 50% prevalence threshold.

**Table 2.**
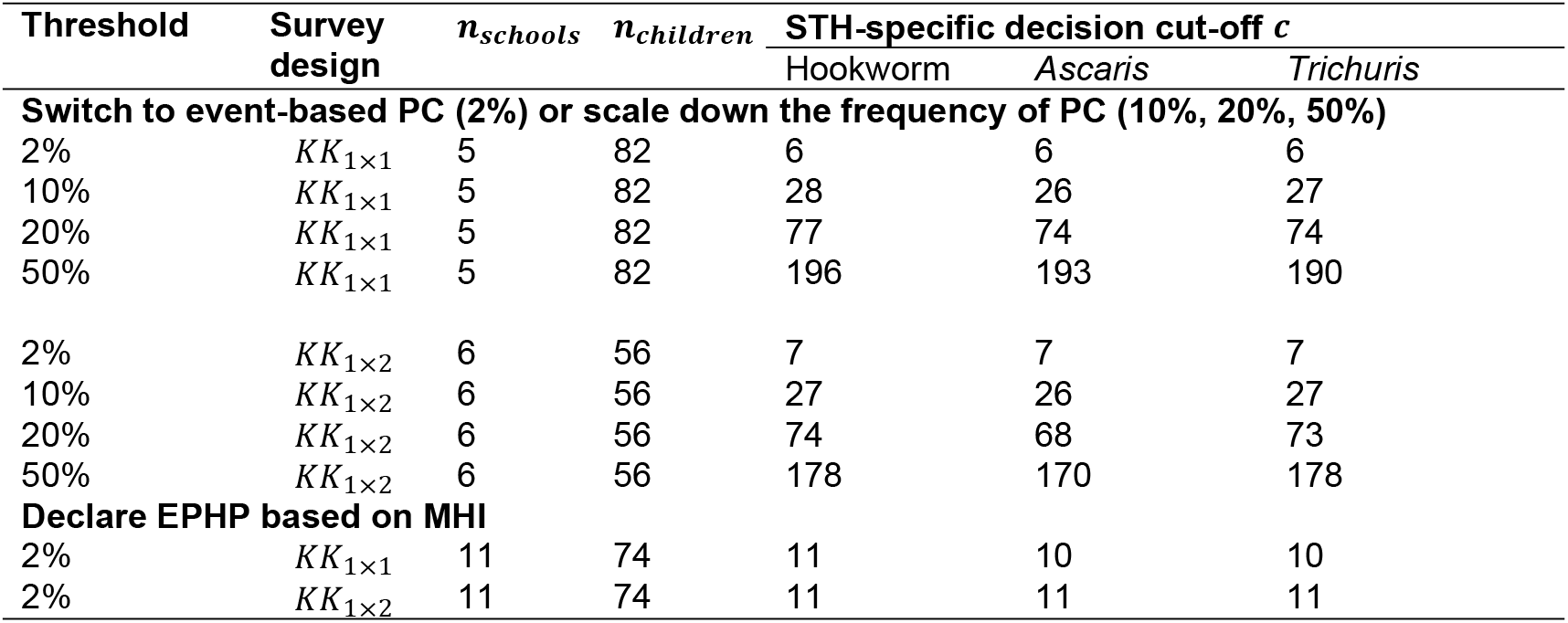
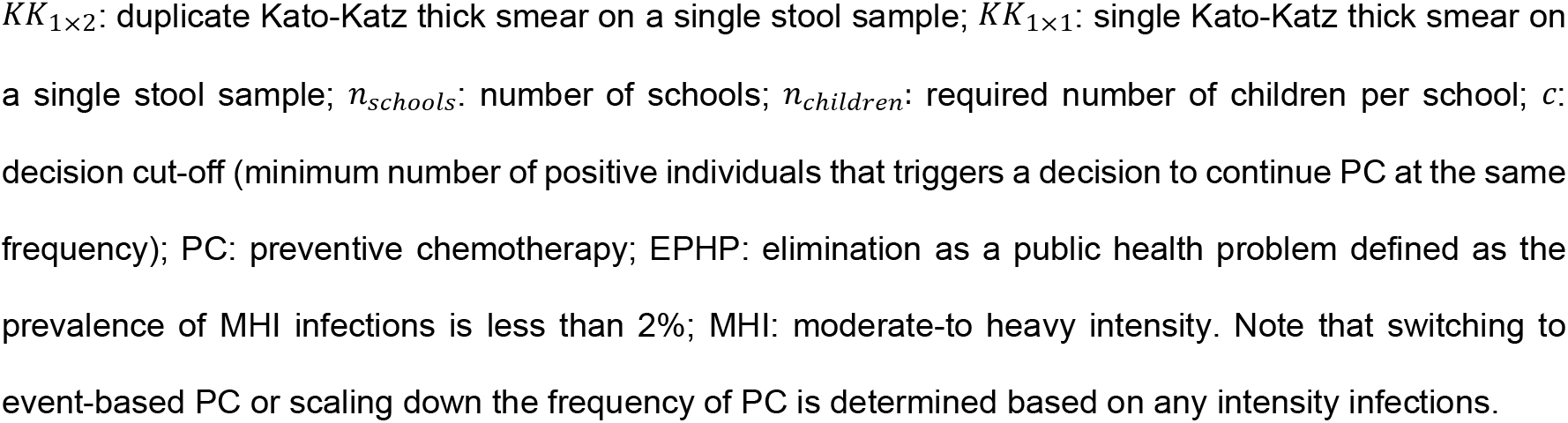
Recommended survey designs for decision making in STH control programs. In this table, we fixed the maximum risk of undertreatment at 1% and overtreatment at 20% for switching to an event-based PC, scaling down PC frequency, and declaring EPHP. The complete set of prior means and corresponding most conservative degree of certainty *κ* to switch to an event-based PC (or declare EPHP) is presented in **Table S3**.

| Threshold | Survey design | $n_{schools}$ | $n_{children}$ | STH-specific decision cut-off $c$ | | |
| --- | --- | --- | --- | --- | --- | --- |
|  |  |  |  | Hookworm | *Ascaris* | *Trichuris* |
| Switch to event-based PC (2%) or scale down the frequency of PC (10%, 20%, 50%) |  |  |  |  |  |  |
| 2% | $KK_{1 \times 1}$ | 5 | 82 | 6 | 6 | 6 |
| 10% | $KK_{1 \times 1}$ | 5 | 82 | 28 | 26 | 27 |
| 20% | $KK_{1 \times 1}$ | 5 | 82 | 77 | 74 | 74 |
| 50% | $KK_{1 \times 1}$ | 5 | 82 | 196 | 193 | 190 |
| 2% | $KK_{1 \times 2}$ | 6 | 56 | 7 | 7 | 7 |
| 10% | $KK_{1 \times 2}$ | 6 | 56 | 27 | 26 | 27 |
| 20% | $KK_{1 \times 2}$ | 6 | 56 | 74 | 68 | 73 |
| 50% | $KK_{1 \times 2}$ | 6 | 56 | 178 | 170 | 178 |
| Declare EPHP based on MHI |  |  |  |  |  |  |
| 2% | $KK_{1 \times 1}$ | 11 | 74 | 11 | 10 | 10 |
| 2% | $KK_{1 \times 2}$ | 11 | 74 | 11 | 11 | 11 |

*KK*_1×2_: duplicate Kato-Katz thick smear on a single stool sample; *KK*_1×1_: single Kato-Katz thick smear on a single stool sample; *n*_*schools*_: number of schools; *n*_*children*_: required number of children per school; *c*: decision cut-off (minimum number of positive individuals that triggers a decision to continue PC at the same frequency); PC: preventive chemotherapy; EPHP: elimination as a public health problem defined as the prevalence of MHI infections is less than 2%; MHI: moderate-to heavy intensity. Note that switching to event-based PC or scaling down the frequency of PC is determined based on any intensity infections.

## Discussion

To further support the efforts of STH control programs, we revised our existing LQAS framework, aiming to make the interpretation of the risks of incorrect decisions more intuitive for programs managers. Using this revised framework, we determined the most cost-efficient survey design for decision-making in STH control programs with regard to a shift towards event-based deworming, declaring EPHP and scaling down PC.

### An intuitive risk-based sampling framework for STH control programs

In our previous framework, the risks of incorrect decision-making were previously conditioned on two predefined values of true underlying prevalence (lower and upper limits of the grey zone) as described elsewhere [14] and illustrated in **Fig 1A**, making its interpretation challenging and not straightforward. Our aim was to make the interpretation of the risk of incorrect decision-making more intuitive while providing the opportunity to further customize the survey design to local settings. Although our revised framework allows for a more intuitive interpretation of the risk of incorrect decision-making, four required parameters were still needed, most notably the maximum acceptable risk of overtreatment and undertreatment, the prior mean *µ*, and the degree of certainty *κ*. To address this, we determined for each prior mean *µ*, the most conservative degree of certainty *κ* (**Table S3**). Accounting for prior knowledge regarding expected true prevalence is more realistic because M&E of STH control programs is performed after several rounds of PC (i.e., 5 to 6 years [7]). Thus, program managers likely have information regarding the expected prevalence based on historical survey data, routine surveillance, or programmatic data within their implementation units. However, translating these expectations into a prior mean as required by this current framework can still be challenging.

It is clear that choices of mean prior and degree of certainty have an impact on the sample size and thus the total survey costs (**Figs 3, S1 and S5**). The association between the degree of certainty *κ* and the total survey cost for a given prior mean *µ* was concave. At lower □ values (e.g., 100), beta distribution becomes J-shaped, with most probability mass near zero, reflecting strong prior belief that prevalence is very low and requires small sample size, and thus results in low total survey cost. At moderate □ values (e.g., 200), the distribution becomes bell-shaped with probability mass spread below and above the prior mean *µ*, indicating greater uncertainty and therefore requiring a large sample size. In contrast, at high value of degree of certainty □, the distribution becomes narrow and sharply peaked around the mean, implying strong belief that the prevalence is low and below the decision threshold, and requires a relatively small sample size. To ensure that the sample size is sufficient for reliable decision-making, we propose considering the most conservative prior certainty level □ which requires the largest sample size (see **Table S3**). Moreover, when the prior mean *µ* is set close to the prevalence threshold, the required sample size and total survey cost size markedly increase. This highlights the importance of carefully choosing a reasonable prior mean *µ* to balance precision with feasibility.

### Agreeing on the acceptable level of risk of incorrect decisions

Reducing maximum acceptable risk of incorrect decision-making clearly leads to an increase in the total survey cost. We considered a maximum risk of undertreatment of 1% to be reasonable and fixed the allowable risk of overtreatment to 20%. However, consensus within the STH community is needed on what constitutes an acceptable level of risk of overtreatment and undertreatment. This is because these risks can vary across program decisions. The relevance of these risks lies in their programmatic consequences, particularly the risk of undertreatment, which can lead to re-emergence of STH morbidity and loss of prior investment in control efforts. Because of these potential consequences, the maximum acceptable risk of undertreatment must be kept low, which directly translates into larger required sample size and higher total survey cost. Therefore, we recommend further studies to elicit acceptable risk values from the wider STH community, including but not limited to policymakers, implementers (e.g., NGOs), and scientists. To this end, we recommend using the IDEA (investigate, discuss, estimate, and aggregate) protocol for structured expert judgment [15]. This protocol can be employed both locally (face-to-face meetings) and remotely. A similar process would be required to apply this framework to other NTDs.

### Survey design choices around the number of Kato-Katz thick smears

We conclude that deploying duplicate KK thick smears on one stool sample was the preferred survey design when making decisions regarding switching to an event-based PC. Although this approach increases the total survey cost by approximately 10% compared to single KK (**Table 1**), it provides the added benefit of internal quality control, improving diagnostic reliability. Similarly, for declaring EPHP, deploying duplicate KK results in a more modest 8% increase in the total survey cost (**Table S4**), further supporting its use as the preferred and most cost-efficient survey design. Our preferred choice of deploying duplicate KK aligns with previous findings for switching to event-based PC but differs for declaring EPHP, where our previous study favoured single KK [11]. This difference is explained by the fact that previous analyses did not incorporate the benefits of the possibility of internal quality control. In addition, the required sample size to declare EPHP was approximately 35% lower than reported in the previous framework [11]. This can be explained by the distinct conceptual approaches of the two frameworks, and we additionally corrected for scale differences, as the school-level mean was expressed in EPG.

### Recommendations to monitor and evaluate STH control programs

We recommend deploying duplicate KK on one stool sample for switching to an event-based PC or scaling down PC for declaring EPHP. These results are particularly welcome since the WHO recently released its M&E framework manual to support schistosomiasis and STH control programs [6]. In this manual, the WHO described how to perform impact assessment using various methods such as cluster surveys, LQAS, and model-based geostatistical approaches. In this manual, the WHO recommends LQAS approaches to sample 332 children with the corresponding decision cut-off for 2%, 10%, 20%, and 50% prevalence thresholds. These decision cut-offs regarding any intensity of infection were defined as follows: zero for below 2%, 1-20 for 2-10%, 21-48 for 10-20%, 49-144 for 20-50% and 145 or higher for prevalence greater or equal to 50% [6]. It is important to note that the method used to derive this sample size and decision cut-off was not explicitly described in the manual, making it difficult to assess the assumptions underlying their calculation. In addition, the sample size and associated cut-offs differ from those identified in our current framework, which explicitly incorporates risk and quality control considerations. Finally, any recommendations to declare EPHP are still missing in the WHO manual.

To further support programs manager, we identified the recommended survey designs when deploying duplicate KK and for informing decisions on switching to event-based PC or scaling down PC frequency (**Table S5**). For this, we evaluated two prior mean assumptions (1% and 1.4%) and two options of certainty (high *vs*. low). Under a 1% prior mean, the required sample size was 6 schools with 56 children per school for the high certainty option, compared with 3 schools with 30 children per school for the lower-certainty option. When fixing the prior mean at 1.4%, sample size requirements increased to 16 schools (100 children per school) for high certainty and 10 schools (68 children per school) for lower certainty (**Table S5**).

Besides the LQAS approach, model-based geostatistics has also been shown to be cost-efficient for M&E of STH control programs [16–19]. Yet, the adoption of this method requires additional skills in geostatistical analysis. The fundamental difference between our LQAS approach and model-based geostatistics is the site selection process (e.g., schools). While the LQAS approach uses a simple random sampling method, the geostatistical approach adopts a spatially regulated sampling approach. Given that using simple random sampling to select sites may result in sampling near-neighboring schools that partially duplicate each other’s information. Therefore, we recommend using the LQAS approach together with a spatially regulated sampling approach for the site selection. Further research could focus on how this integration of spatially regulated sampling would affect the program decision-making based on the survey designs recommended in the current study.

### Limitations of our study

It is important to note that the estimated degree of variability in egg counts across schools (**Table S1**) was based on data obtained from a school-based survey carried out in Ethiopia [12], assuming an implementation unit size of approximately 130,000 inhabitants. Therefore, in settings where the implementation unit size is significantly larger, we recommend assessing at the sub-implementation unit level to minimize the risk of prematurely switching to event-based PC or undertreatment. This risk arises because larger implementation units often encompass areas with heterogeneous transmission patterns, where combining data across diverse schools may lead to inappropriate decisions. Moreover, the total survey costs estimates were also based on the Ethiopian context, and our results were restricted to certain values of prior knowledge and risk of incorrect decision making. A key strength of this study, however, is that the database of simulations was built independently of any specific epidemiological (prior knowledge) or operational context (total survey cost), making it setting-agnostic. As such, we can easily adapt the framework output to specific contexts by incorporating context-specific prior information and cost data without having to rerun the database of simulations, thereby enabling efficient setting-specific decision-making.

## Conclusion

We developed an intuitive sampling framework for setting-specific decision-making in STH control programs. We identified the most cost-efficient survey designs for most important program decisions for a set of somewhat subjective but reasonable choices regarding acceptable levels of risk of incorrect decision making. Reaching consensus within the STH community on this acceptable level of risks is crucial to support evidence-based decision-making. In addition, further research could focus on how spatially regulated sampling can further improve the accuracy and cost-efficiency of LQAS survey designs.

## Data Availability

All relevant data are within the manuscript and its supporting information files

## Supporting information captions

**Fig S1.**
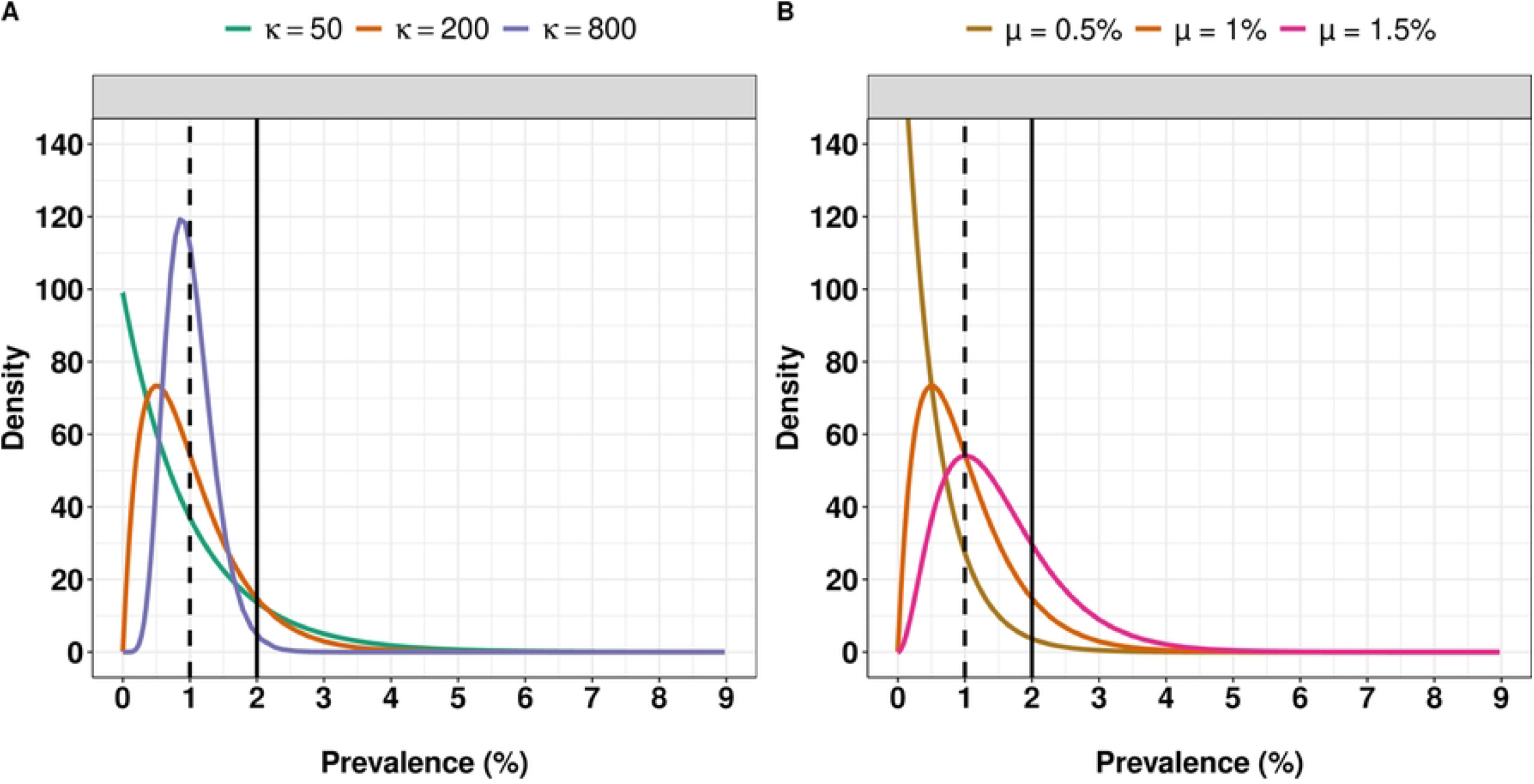
Impact of prior mean and degree of certainty of the shape of beta distribution around the 2% program prevalence threshold. To do this, we first set the mean *μ* to 1% in **Panel A** while varying the degree of certainty □ to 50, 200 and 800. In **Panel B**, we set the mean of degree of certainty to 200, and the mean *μ* values of 0.5%, 1% and 1.5%. Vertical dotted and solid lines indicate the prior mean of 1% and the prevalence threshold of 2%, respectively.

**Table S1. Parametrisation of the simulation framework for various sources of variability in Kato-Katz thick smear egg counts.** We parametrized the lognormal distribution of the variability in mean eggs per gram of stool (EPG) across schools using the mean and standard deviation on the logarithmic scale. For the gamma distribution of the inter-individual variability and day-to-day variability in mean EPG, we used the shape parameter *k* and scale 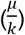, where *μ* is the distribution’s mean. Note that lower values of *k* mean more heterogeneity and *β*_1_ was divided by 24 to correct for the scale difference, as the school-level mean is expressed in EPG.

**Fig S2.**
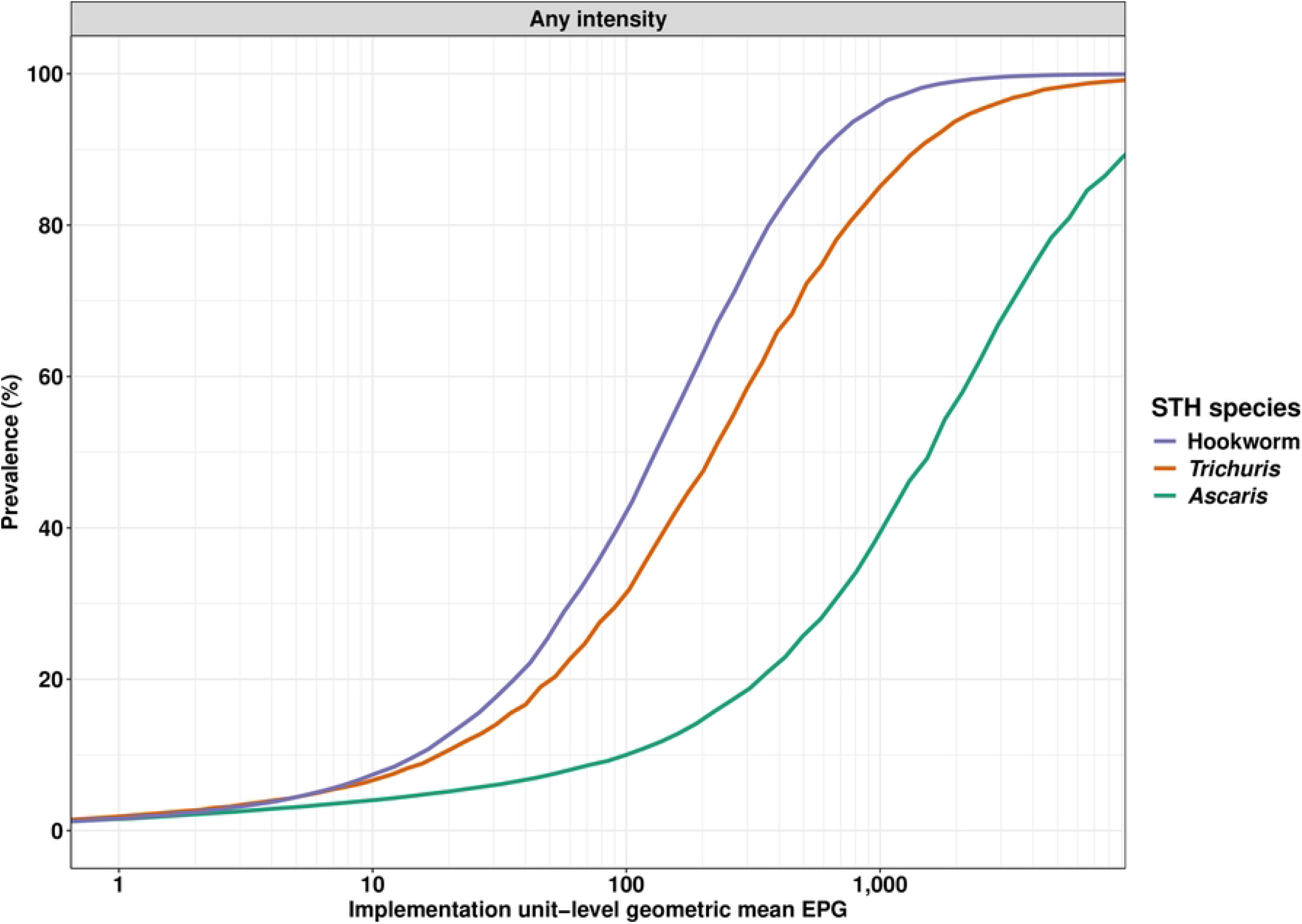
Association between implementation unit-level geometric mean EPG and the resulting true prevalence of any intensity across three STH species. For this, we first determined true prevalence at the implementation unit level when testing 3,000 schools and recruiting 6,000 per school for every mean number of eggs per gram of stool (EPG) when deploying single Kato-Katz thick smear on a single stool sample, accounting for the different sources of variability in egg counts (**Table S1**). Note that the logarithm of these mean EPGs was used in our simulation framework.

**Fig S3.**
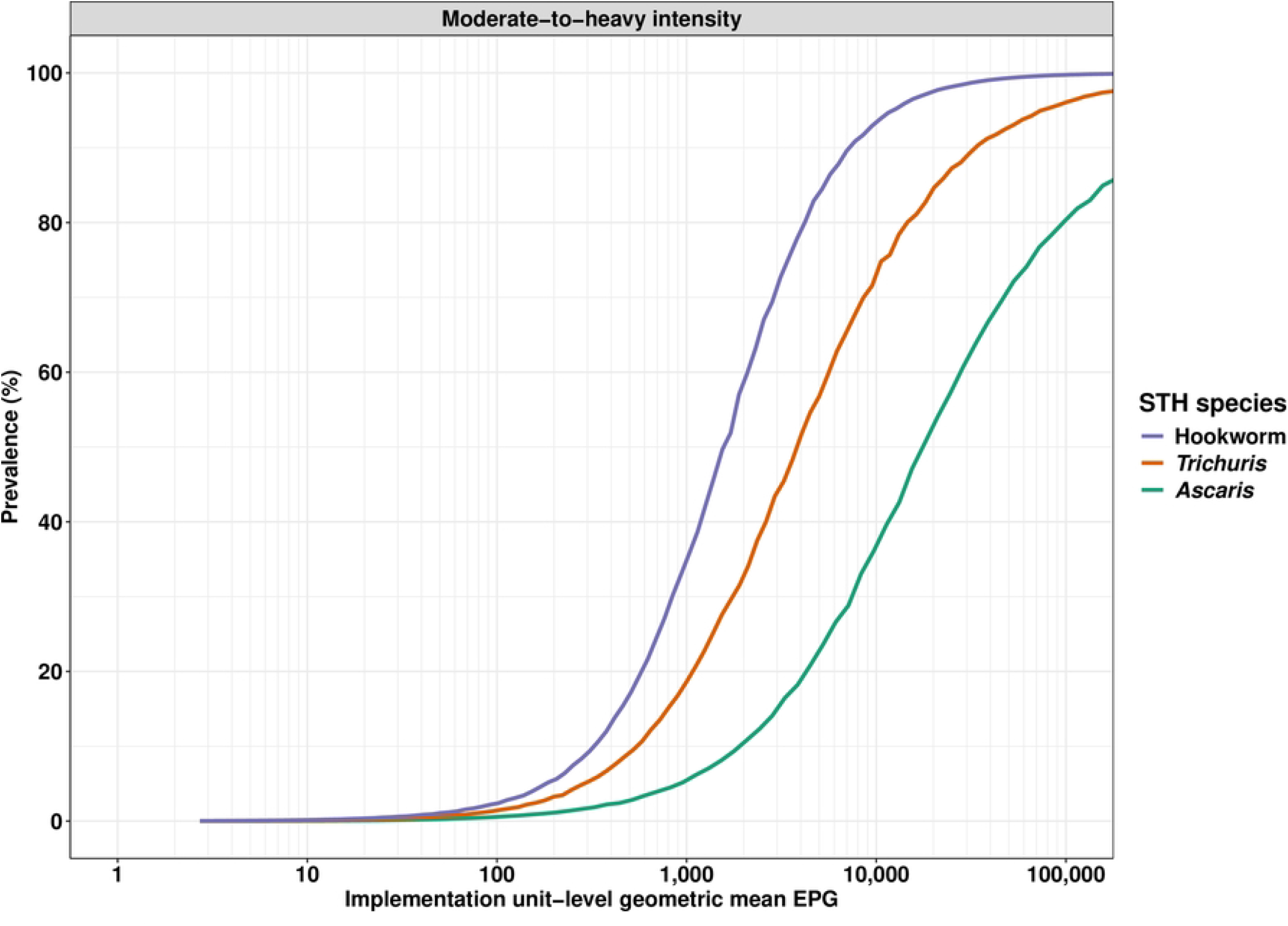
Association between implementation unit-level geometric mean EPG and the resulting true prevalence of moderate-to-heavy intensity infections across three STH species. To do this, we first determined true prevalence at the implementation unit level when testing 3,000 schools and recruiting 6,000 children per school for every mean number of eggs per gram of stool (EPG) when deploying single Kato-Katz thick smear on a single stool sample, accounting for the different sources of variability in egg counts (**Table S1**). We used the WHO classification of moderate-to-heavy intensity infections (MHI), which is 5,000 EPG for *Ascaris*, 2,000 EPG for hookworm, and 1,000 EPG for *Trichuris*. In our simulation, the logarithm of these mean EPG was used.

**Table S2. Overview of the cost parameters when deploying KK testing in an Ethiopian setting**

**Table S3. Prior mean and corresponding most conservative degree of certainty when determining survey design to scale down or declare EPHP.** The most conservative *κ* corresponds to the one *κ* that yields the largest sample size.

**Fig S4.**
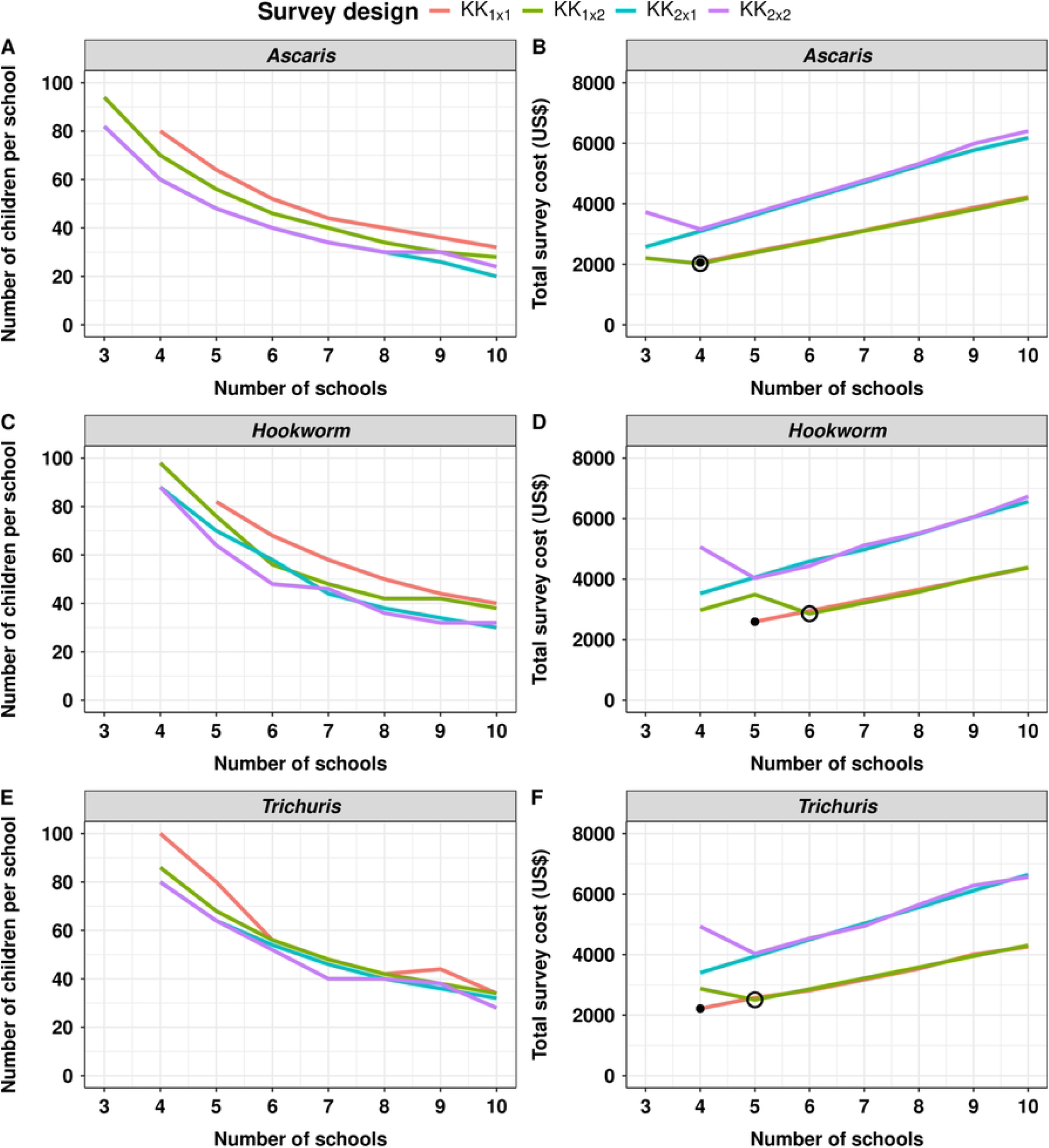
The identification of the most cost-efficient survey design to reliably switch to an event-based PC. This figure illustrates the required number of children per school (*n*_*children*_; **Panels A, C** and **E**) and the corresponding total survey cost (*C*_*tot*_; **Panels B, D** and **F**) as a function of the number of sampled schools (*n*_*schools*_) for different survey designs and soil-transmitted helminths species (*Ascaris*: **Panels A** and **B**; hookworm: **Panels C** and **D**; *Trichuris*: **Panels E** and **F**). The survey designs (*KK*_*a* × *b*_) varied in the number of stool samples per child (= *a*) and the number of Kato-Katz thick smears per sample (= *b*). The black bullet point in Panels B, D and F indicates the most cost-efficient survey design. In other words, the number of schools that minimizes the costs while ensuring reliable decision-making. The open circle indicates the most cost-efficient survey design when preparing two slides per sample, which automatically adds free quality control to the survey design.

**Fig S5.**
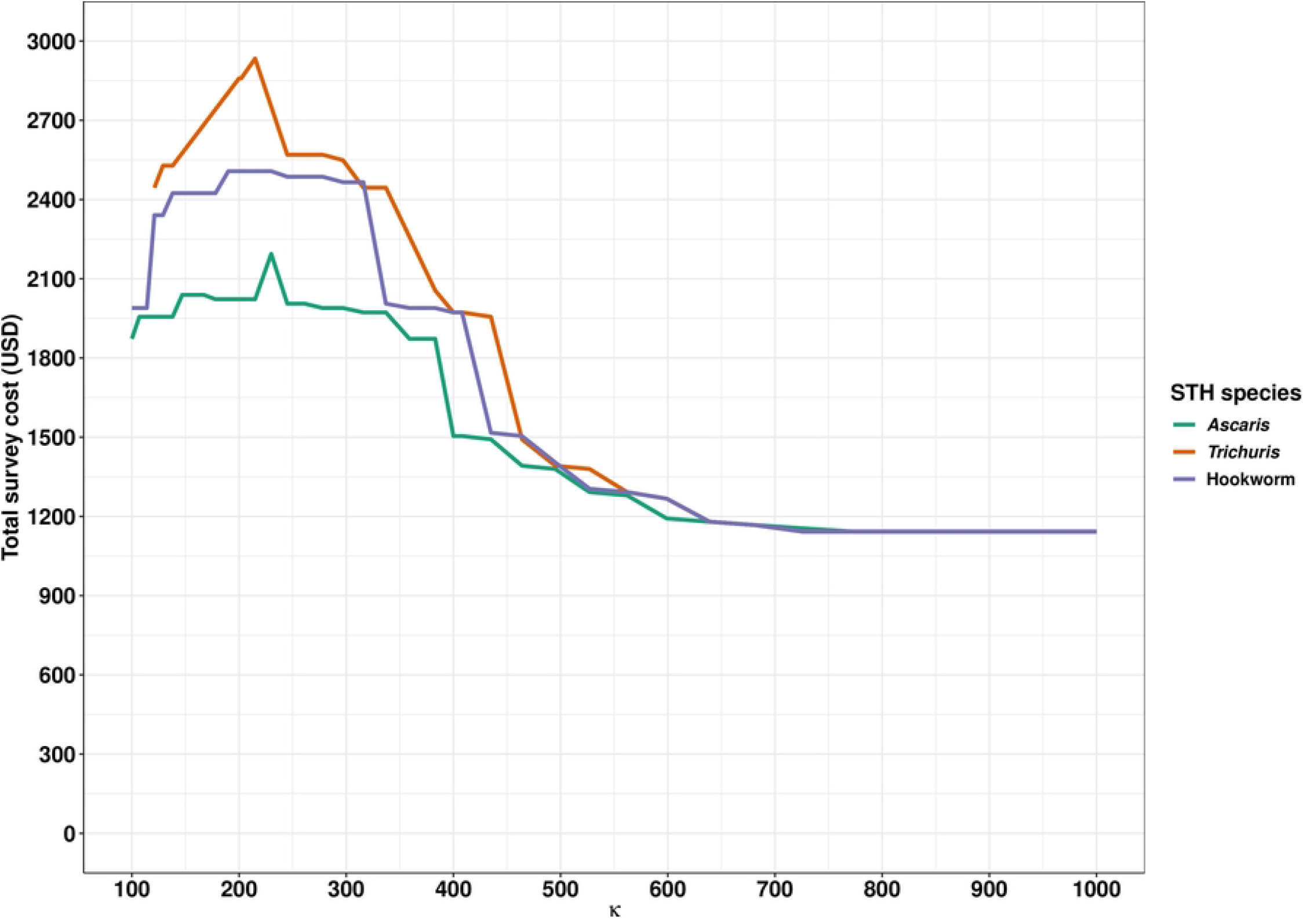
The required total survey cost to reliably switch to an event-based preventive chemotherapy as a function of the degree of certainty expressed by *κ*. This figure presents the required total survey cost for switching to an event-based PC for each STH species as a function of the degree of certainty (*κ*). For this, we set the prior mean to 1% and the risk of incorrect decisions to 20% for overtreating and 1% for the risk of undertreating.

**Table S4. The cost-efficient survey design to declare EPHP.** This table represents the required sample size (*n*_*schools*_ × *n*_*children*_), the decision cut-off *c* and the total survey cost (*C*_*tot*_) for surveys based on screening one stool sample with a single Kato-Katz thick smear (*KK*_1×1_). Note that we assumed the maximum number of children per school. Also, we set the risk of undertreating to 1% while fixing the maximum allowed the risk of overtreatment was 20%, a certainty level □ of 200 and prior mean of 1%.

**Table S5. Recommended survey designs for decision making in STH control programs.** In this table, we fixed the maximum risk of undertreatment at 1% and overtreatment at 20% for switching to an event-based PC or scaling down PC frequency. To this end, we first identified the required sample size to switch to event-based PC when considering a prior mean of 1% and 1.4% and two options for the degree of certainty: high (200) and low (800). We further determined the corresponding decision for scaling down the PC frequency while using the required sample for 2% threshold and their corresponding prior mean.

